# Could sensorimotor factors in acute low back pain explain long-term pain and disability? A secondary analysis of longitudinal data from a randomised controlled trial

**DOI:** 10.64898/2026.07.28.26359143

**Authors:** Claudia Côté-Picard, Jean-Sébastien Roy, Hugo Massé-Alarie

## Abstract

Treatments for chronic low back pain (LBP) provide only small improvements in pain and disability compared with placebo. Targeting mechanisms involved in the persistence of pain and disability following an episode of acute LBP (ALBP) may enhance treatment effectiveness. Sensorimotor alterations have been observed in individuals with chronic LBP (CLBP), but their role in the development of CLBP remains unclear. In this secondary analysis of longitudinal data from 99 participants with ALBP, the causal associations between pain sensitivity and erector spinae muscle activation measured in the acute phase of LBP and pain and disability at 6- and 12- month follow-ups were explored. Negative binomial regressions revealed that greater lumbar muscle activation at baseline was associated with lower disability at 6 months (incidence rate ratio [IRR] 0.33 [95% CI 0.13 to 0.85], p=0.02). Higher lumbar pressure pain threshold was also associated with lower disability at 6 months (IRR 0.87 [95% CI 0.79 to 0.97], p=0.009). At 12 months, greater lumbar and thoracic muscle activation were associated with lower disability (IRR 0.11 [95% CI 0.03 to 0.35], p<0.001 and IRR 0.09 [95% CI 0.02 to 0.33], p<0.001, respectively). The findings suggest that increased thoracolumbar muscle activation during the acute phase may act as a protective mechanism and support recovery. However, the association with pain sensitivity should be interpreted with caution, as only one of 16 models reached statistical significance. Longitudinal studies assessing the evolution of these sensorimotor factors could improve the understanding of their contribution in the development of CLBP.

**Perspective:** This article presents exploratory analyses of causal associations between sensorimotor outcomes in acute low back pain and levels of pain and disability in the long-term. Increased muscle activation explained better long-term outcomes, whereas increased pain sensitivity explained poorer long-term outcomes.

## INTRODUCTION

Chronic low back pain (CLBP) places a substantial burden on individuals and society and is the leading cause of years lived with disability globally.^1^ Acute low back pain (ALBP) generally improves rapidly within six weeks of onset (e.g., 30% and 21% decrease in pain and disability levels, respectively). However, although recovery rates vary greatly between studies,^2^ a 2013 meta-analysis reported that low back pain (LBP) persists in 57% (95% CI 46–68%) of individuals at 6 months and 65% (95% CI 54–75%) at 12 months following acute or subacute LBP.^3^ Treatments for CLBP provide only small improvements in symptoms compared with placebo.^1,4,5^ Targeting mechanisms involved in the persistence of pain and disability following an episode of ALBP might therefore enhance treatment effectiveness.^6^ Healthcare professionals commonly use interventions targeting sensorimotor components,^7,8^ given evidence that these are altered in individuals with LBP.^9–12^ However, to our knowledge, only one longitudinal study thoroughly tested potential causal sensorimotor mechanisms contributing to the development of CLBP.^13^ It showed that lower somatosensory cortex excitability, measured in ALBP, was causally associated with future CLBP, suggesting that this cortical area contributed to the transition from ALBP to CLBP.

A factor that was suggested to contribute to chronic pain development is the long-term persistence of central sensitization,^14^ defined as an increased responsiveness of nociceptive neurons within the central nervous system.^15^ As it is impossible to directly measure this construct in humans, the term Human Assumed Central Sensitization (HACS) was recently introduced to characterize the presence of clinical signs consistent with central sensitization (e.g., as captured by quantitative sensory testing [QST]).^16^ HACS has been observed in acute, subacute and chronic LBP,^12,17^ with higher effect sizes in CLBP than ALBP compared to pain-free controls.^17^ Its role in predicting the transition from acute to chronic LBP remains however unclear. Indeed, a 2016 meta-analysis reported inconclusive results regarding the prognostic value of QST in LBP,^18^ and recent prospective cohort studies have shown that QST measures in ALBP did not predict chronic pain or recovery.^19–21^ In contrast, a 2018 longitudinal study reported that HACS during the subacute phase of LBP was associated with chronic pain development.^22^ Furthermore, a 2019 meta-analysis from broader musculoskeletal pain populations showed that HACS was associated with worse pain and disability at follow-up.^23^ However, existing studies rarely controlled rigorously for potential confounders, limiting causal inference. As such, it remains unclear whether these associations reflect true causal mechanisms underlying the development of CLBP.

Back muscle adaptations could also be implicated in CLBP development. The “motor adaptation to pain” theory proposes that acute or experimentally induced pain leads to modifications in motor control aimed at protecting the injured/threatened tissue in the short-term.^24^ There is a lack of empirical data supporting this theory in patients with clinical ALBP, but increased lumbar erector spinae muscle activation during a trunk flexion is observed in more than half of patients with CLBP.^25^ If higher erector spinae activation in ALBP reflects such protective response, it would be expected to be associated with better long-term recovery, but this is currently unknown. Moreover, persisting muscle hyperactivation may be detrimental, as it increases the load on lumbar spine structures and limits movement.^24^ Overall, it remains unclear if higher or lower erector spinae muscle activation during ALBP is associated with the persistence of symptoms.

The aim of this study was to explore the causal associations between pain sensitivity and erector spinae muscle activation in ALBP and pain and disability at 6- and 12-month follow-ups. We hypothesized that initial pain hypersensitivity – suggesting HACS – and higher erector spinae muscle activation would be associated with greater pain and disability at both follow-ups.

## METHODS

### Study design and setting

This study is a secondary analysis of longitudinal data collected within a randomised controlled trial involving individuals with ALBP. This trial was registered on Clinicaltrials.gov (NCT03986047) and approved by the Research Ethics Committee of the CIUSSS de la Capitale-Nationale (#2019-1731, RIS – CER CIUSSS-CN). Participants provided written informed consent before enrolling in the study. There was no public involvement in the study’s conceptualization, design and conduct. The trial protocol and results were previously published.^26,27^ Recruitment took place from April 2019 to March 2023.

Three different interventions (heatwrap and exercise, heatwrap alone, and sham heatwrap) were tested in the original trial, with no between-group difference observed for primary and secondary outcomes.^26,27^ Therefore, participants from all three groups were pooled and analysed as a single cohort. We followed the Strengthening the Reporting of Observational Studies in Epidemiology (STROBE) guidelines to report the findings of the current study.^28^ Baseline assessments were conducted at the Center for Interdisciplinary Research in Rehabilitation and Social Integration (Cirris), Québec City, Canada. Measurements of QST and back muscles’ activity via electromyography were taken by an assessor blinded to the group allocation, who was a physiotherapist trained in QST and electromyography. Participants completed online questionnaires on demographic characteristics at baseline and symptoms at baseline, 6 months and 12 months.

### Study population

Participants were recruited through e-mail listings at Université Laval, the Quebec Back Pain Consortium database^29^ and social media ads. We included participants aged 18 to 65 years with ALBP (pain in the lumbar region, between the 12^th^ rib and the gluteal fold, for six weeks or less with or without leg pain)^30^ who understood written French. Participants had to have experienced limited daily life activities for more than one day and a minimal score of 10/100 to the Oswestry Disability index (ODI).^31^ Exclusion criteria were LBP in the 3 months preceding the current episode, signs of a serious pathology or neural impairment, pregnancy, important skin lesion in the lumbar area or cognitive impairments. Other exclusion criteria were a history of a spinal surgery, a recent change in medication that could influence pain (e.g., psychotropic drugs) or corticosteroid injection in the 6 weeks preceding the enrolment in the study.

### Outcomes

Outcomes were the average level of pain over the past week, measured on a 0 (no pain) to 10 (worst imaginable pain) numerical pain rating scale (NPRS),^32^ and the level of disability measured with the ODI (0 to 100 scale, where a higher score reflects more disability).^31^ Both outcomes were measured at 6- and 12-months follow-ups. These questionnaires were chosen as they both have strong construct validity, and excellent test-retest reliability in LBP populations.^31,33–35^ They are also considered by experts as core outcome measurements in clinical trials for non-specific LBP.^36^

### Exposure variables – pain sensitivity

Pressure pain threshold (PPT) and temporal summation of pain (TSP) were used to quantify local and remote pain sensitivity at baseline. Local sensitivity may reflect both peripheral and central sensitization, whereas remote sensitivity would be indicative of central sensitization.^37^ Static sensory testing, as PPT, reflects the basal state of the nociceptive system, and when tested at the site of pain, may predominantly reflect peripheral pain sensitization.^23,38^ In contrast, TSP is a test of central integration of pain, including descending pain modulation, and suggests the presence of central sensitization when enhanced.^38^ Two different QST measures were chosen as it was suggested that varied modalities give more information on the pain modulation system.^39^ Standardized instructions freely translated to French from the German Research Network on Neuropathic pain (DFNS) were given to participants prior testing.^40^ There was a familiarisation period where PPT was applied on wrist extensors and TSP on the dorsum of the hand. Both measures have excellent within-session reliability in people with CLBP.^41^ The testing sequence was randomised by a computer random number generator to control for the potential effect of testing order.

#### Pressure pain threshold (PPT)

PPT was measured close to the site of pain (lumbar region) and remotely (contralateral leg) with a handheld digital algometer (1cm^2^ probe–FPIX, Wagner Instruments, Greenwich, USA). The first PPT was measured on the lumbar erector spinae muscles 3 cm lateral to the L4-L5 joint on the most painful side, or on the right side if the pain was bilateral. The second one was measured on the tibialis anterior (TA) contralateral to the most painful side. Pressure was applied at a rate of ∼0.5kg/cm^2^/s, and 3 consecutive trials were performed with a 30-second pause between measurements. The measure of PPT was obtained when participants indicated that the pressure became painful. The mean of 3 trials was calculated to obtain the PPT outcome at each site.

#### Temporal summation of pain (TSP)

As for PPT, two measures of TSP were used: site of pain (lumbar region) and remote (contralateral foot). TSP was tested with a Pinprick stimulator (256 mN, MRC Systems GmbH) at the L4-L5 intervertebral joint line and on the 3^rd^ cuneiform bone on the dorsal aspect of the foot contralateral to the most painful side. A single stimulus was applied to the tested site, then 10 stimuli were applied at a frequency of 1 Hz monitored by a flashing metronome out of the participant’s sight. The TSP measure was the difference of the greatest pain intensity through the 10 stimuli and the pain intensity during a single stimulus on the NPRS.^42^ This procedure was performed 3 times with a 30-second break between measurements, and the mean of 3 trials was calculated to obtain the TSP outcome at each site.

### Exposure variables – erector spinae muscle activation

Wireless surface electromyography (sEMG) sensors (TrignoTM Wireless System, Delsys) were used to record muscle activation at T12 and L5 erector spinae at a sampling rate of 1000 Hz during a trunk flexion task. The sensors were placed, following SENIAM recommendations, on the most painful side of the back or at the right side if the pain was bilateral.^43^ The sensors included inertial measurement units (IMU) that sampled the 3-dimension acceleration at 148 Hz. The skin was shaved and cleaned with an alcohol swab. Participants were asked to stand still with feet at shoulder width for 3 seconds, bend the trunk forward without bending the knees to reach full trunk flexion over 5 seconds, remain in full flexion for 3 seconds and return to the initial position over 5 seconds. This procedure was repeated 3 times, with a 30-second interval between measurements.

Raw sEMG data from both sensors were imported into MATLAB R2019a (The Mathworks inc., Natick, Massachusetts, USA), band-pass filtered using a 4^th^ order Butterworth filter (20-450 Hz) and full-wave rectified. The EMG signal was illustrated as a moving average in a 250 ms sliding window. Raw sagittal plane acceleration data were also imported in MATLAB and resampled to match sEMG sampling frequency (1000Hz). A custom graphical user interface was used to superimpose the raw acceleration signal to the rectified and filtered sEMG signal and used to identify each phase of the movement: (i) standing, (ii) flexion, (iii) full flexion, and (iv) extension. Peak muscle activation was then identified in each phase. This procedure was repeated for each trial. Peak values obtained during the full flexion and extension phases were averaged through the 3 trials to calculate ratios at each sensor (L5 and T12). The flexion-relaxation ratio (FRR) was defined as the ratio of the mean peak activation during full flexion to that during extension.^25^ FRR has moderate to high test-retest reliability.^44^

### Confounders

We were interested in exploring the total effect of exposures over outcomes, which includes the direct effect of the exposure on the outcome and its indirect effect through a mediating variable.^45^ To estimate such effect, only confounders, i.e. variables that influence both the exposure and outcome, should be controlled in statistical analyses.^46,47^

Directed Acyclic Graphs (DAGs) were used via the DAGitty web application to illustrate potential causal pathways between covariates, exposures and outcomes. DAGs provide a non-parametric framework to identify the minimal sufficient adjustment set (MSAS) to obtain valid causal effect estimates.^48,49^ Independent DAGs were built for each group of exposure (i.e. one for erector spinae muscle activation [lumbar and thoracic FRR] and one for pain sensitivity [local and remote PPT and TSP]). The outcome was considered as pain intensity or disability level in the long-term (6 and 12 months); the same DAG was used for all outcomes. The included covariates were variables that were measured at baseline in the main RCT and that we found evidence that they could have an influence on either the exposure or the outcome.^48^ Variables were presented as nodes, and arrows were drawn between them to illustrate a causal relationship from a variable to another. It is recommended to saturate the DAGs, i.e. to draw an edge between each variable and to assess each edge for causal relationship.^48^ However, considering that relationships between some covariates have never been assessed and are often difficult to hypothesize, only relationships with empirical support or strong theoretical rational were drawn. Also, we ensured that the absence of edges did not influence the MSAS (e.g., by verifying that it did not remove a plausible confounder). Also, we did not include edges if the MSAS was saturated, i.e. if the addition of a given arrow between two variables would not change the MSAS.

The choice to keep an edge and add an arrow between two given variables was based on three causal criteria, i.e. temporality (the exposure has to precede the outcome), face validity (the relationship has to be plausible) and recourse to theory (formal evidence or expert knowledge supports the relationship).^48^ The codes used to build both DAGs are available in **Supplementary file 1**. We compiled the references alongside the design of the studies that guided our assessment of the relationships between variables in **Supplementary file 2**. DAGs are presented in **Figure 1**. For muscle activation models, the MSAS comprised age, sex, BMI, baseline disability (measured with ODI), pain-related fear of movement (measured with the 11-item Tampa scale of Kinesiophobia),^50^ pain catastrophizing (measured with the Pain catastrophizing scale),^51^ and pain sensitivity (lumbar PPT). For pain sensitivity models, the MSAS included age, sex, smoking status, pain-related fear of movement and pain catastrophizing.

**Figure 1.**
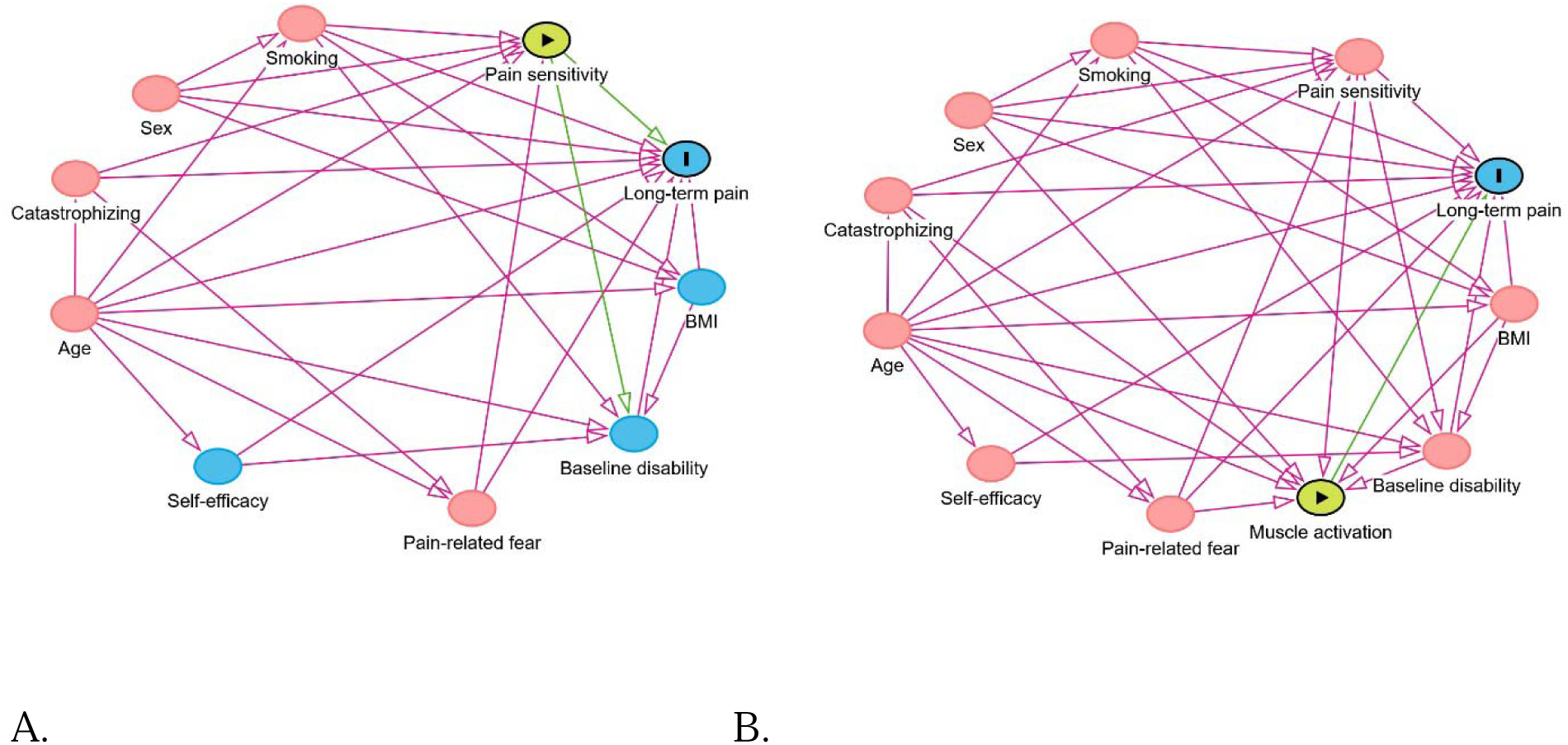
Directed acyclic graph (DAG) for A. Muscle activation and B. Pain sensitivity exposures. Pink nodes are ancestors of exposure and outcome, blue nodes with a black bar are outcomes, green nodes are exposures, and blue nodes are ancestors of outcome. The outcome (long-term pain) represents pain and disability outcomes.

### Statistical analyses

Sample size was calculated for the main RCT.^27,52^ Yet, in the present study, considering an α level of 0.05, a power (1-β) of 0.80, and 5 or 7 predictors, as in our statistical models, at least a small effect size (Cohen’s f^2^=0.09) could be detected with a sample size of 87 participants (**Figure 2**), as calculated with G*Power 3.1.9.7. Statistical analyses were computed using IBM SPSS statistics v.30.0. We set the significance threshold at p<0.05. Baseline demographic and clinical characteristics were presented as counts and percentage for categorical variables and mean and standard deviations (SD) for continuous variables. Correlations between covariates and outcomes were tested using the Pearson coefficient (r) to describe the relationship between potential confounding factors and outcomes and presented in **Supplementary file 3**. These correlation coefficients helped to guide our decisions on keeping or removing an edge between variables included in the DAGs. For causation analyses, we used separate generalized linear models to test the estimated causal effect of each exposure on each outcome, with complete case analyses.^53^ Confounders identified in DAGs (MSAS) for both muscle activation and pain sensitivity exposures were incorporated in the models. Corrections for multiple testing were not used as the study was exploratory.

**Figure 2.**
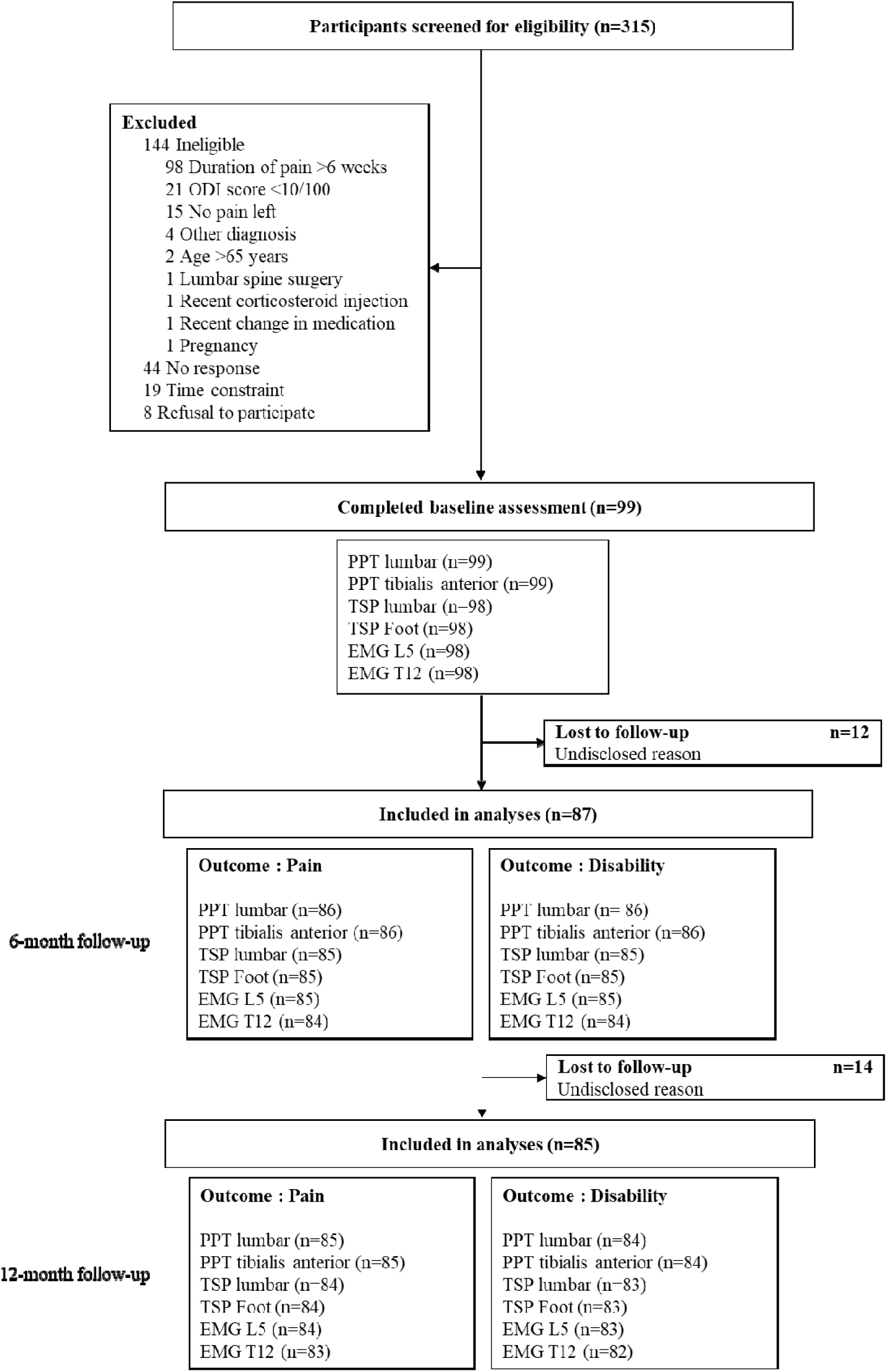
Study flowchart

We first tested the Poisson regression for each model and looked for overdispersion (variance >mean).^54^ There was evidence of overdispersion in all models, so negative binomial regressions were used instead, as recommended.^55^ These models require that the outcome be non-negative integers count data. Our disability variables (ODI at 6 and 12 months) contained non-integers (one question of this tool could be skipped and in this case the result was calculated on a total of 90 points and transformed to percentages), so we rounded the non-integer data to the nearest integer to be able to use generalised linear models. These models provide an Incidence Rate Ratio (IRR), where a value of 1 means no effect of the exposure on the outcome, a value >1 indicates an increase in the outcome with the increase in the exposure, and a value <1 indicates a decrease in the outcome with an increase in the exposure.^56^ We tested for multicollinearity by verifying the Variance Inflation Factor (VIF).

## RESULTS

### Participants and study flow

From April 2019 to March 2023, 315 participants were screened for eligibility. Of those, 144 were ineligible, 44 stopped responding, 19 could not participate due to time constraint, and 8 refused to participate. Ninety-nine participants completed baseline assessment. **Figure 2** depicts the study flowchart with the number of participants included in each analysis. The demographic and baseline clinical characteristics of participants are presented in **Table 1**. Most participants were female (62%), the mean age was 36.4 years (SD 12.8), the mean LBP episode duration was 25.2 days (SD 11.9), the average pain intensity over the last week was 5.1/10 (SD 1.9), and the mean disability level was 22.8% (SD 10.9). Follow-ups at 6 and 12 months were completed by 87 and 85 participants, respectively. Twenty participants had at least one missing data in the exposures, covariates or outcomes. Most were female (65%), the mean age was 35.2 years (SD 12.6), mean LBP episode duration was 24.2 days (SD 12.3), average pain intensity over the previous week at baseline was 5.25/10 (SD 2.0), and mean disability level was 23.5% (SD 9.2).

**Table 1.** Baseline demographic and clinical characteristics of study participants.

| Characteristics of study participants | Total (n=99) |
| --- | --- |
| **Demographic characteristics** |  |
| Sex, n of females (%) | 61 (62) |
| Age (years), mean [SD] | 36.4 [12.8] |
| BMI (kg/m<sup>2</sup>), mean [SD] | 26.7 [5.1] |
| Smoking status, n (%) |  |
| Never smoked | 66 (67) |
| Former smoker | 23 (23) |
| Current smoker | 10 (10) |
| Highest education level completed, n (%) |  |
| High school graduate | 7 (7) |
| Certificate or Diploma | 30 (30) |
| Bachelor's degree or higher | 62 (63) |
| Ethnicity, n (%) |  |
| Métis (Indigenous people in Canada) | 1 (1) |
| White | 74 (75) |
| Chinese | 3 (3) |
| Black | 7 (7) |
| Latin American | 5 (5) |
| Arabic | 11 (11) |
| South-East Asian | 1 (1) |
| Occidental Asian | 1 (1) |

| **Clinical characteristics** |  |
| --- | --- |
| Participants with a history of LBP, n (%) | 63 (64) |
| LBP episode duration (days), mean [SD] | 25.2 [11.9] |
| ODI, mean [SD] | 22.8 [10.9] |
| NPRS, mean [SD] | 5.1 [1.9] |
| TSK-11, mean [SD] | 36.3 [6.8] |
| PCS, mean [SD] | 14.6 [9.4] |
| PPT lumbar (kg/cm<sup>2</sup>), mean [SD] | 4.3 [2.4] |
| PPT tibialis anterior (kg/cm<sup>2</sup>), mean [SD] | 4.9 [2.4] |
| TSP lumbar, mean [SD] | 2.2 [1.3] |
| TSP foot, mean [SD] | 2.3 [1.4] |
| EMG ratio flexion/extension L5, mean [SD] | 0.64 [0.28] |
| EMG ratio flexion/extension T12, mean [SD] | 0.56 [0.25] |
| n, number of participants; LBP, low back pain; 95% CI, 95% confidence interval; BMI, body mass index; NPRS, Numerical Pain Rating Scale; ODI, Oswestry Disability Index; TSK-11, Tampa Scale for Kinesiophobia; PCS, Pain Catastrophizing Scale; PPT, Pain pressure threshold; TSP, Temporal summation of pain; EMG, Electromyography. |  |

### Association estimates of sensorimotor exposure for long-term pain and disability

IRRs and their 95% confidence intervals are presented in **Table 2**. This section will present the results of adjusted models. Lumbar PPT was associated to pain-related disability at 6 months (IRR 0.87 [95% CI 0.79 to 0.97], p=0.009), indicating that an increase in PPT was associated with a decrease in disability level at 6 months. No other significant associations were found for pain sensitivity exposures. Lumbar FRR was associated with disability levels at 6 months (IRR 0.33 [95% CI 0.13 to 0.85], p=0.02) and 12 months (IRR 0.11 [95% CI 0.03 to 0.35], p<0.001). In contrast, thoracic FRR was significantly associated with disability levels only at 12 months (IRR 0.09 [95% CI 0.02 to 0.33], p<0.001). Results for both lumbar and thoracic FRR reflect that high muscle activation of erector spinae was associated with less disability in the long-term.

**Table 2.**
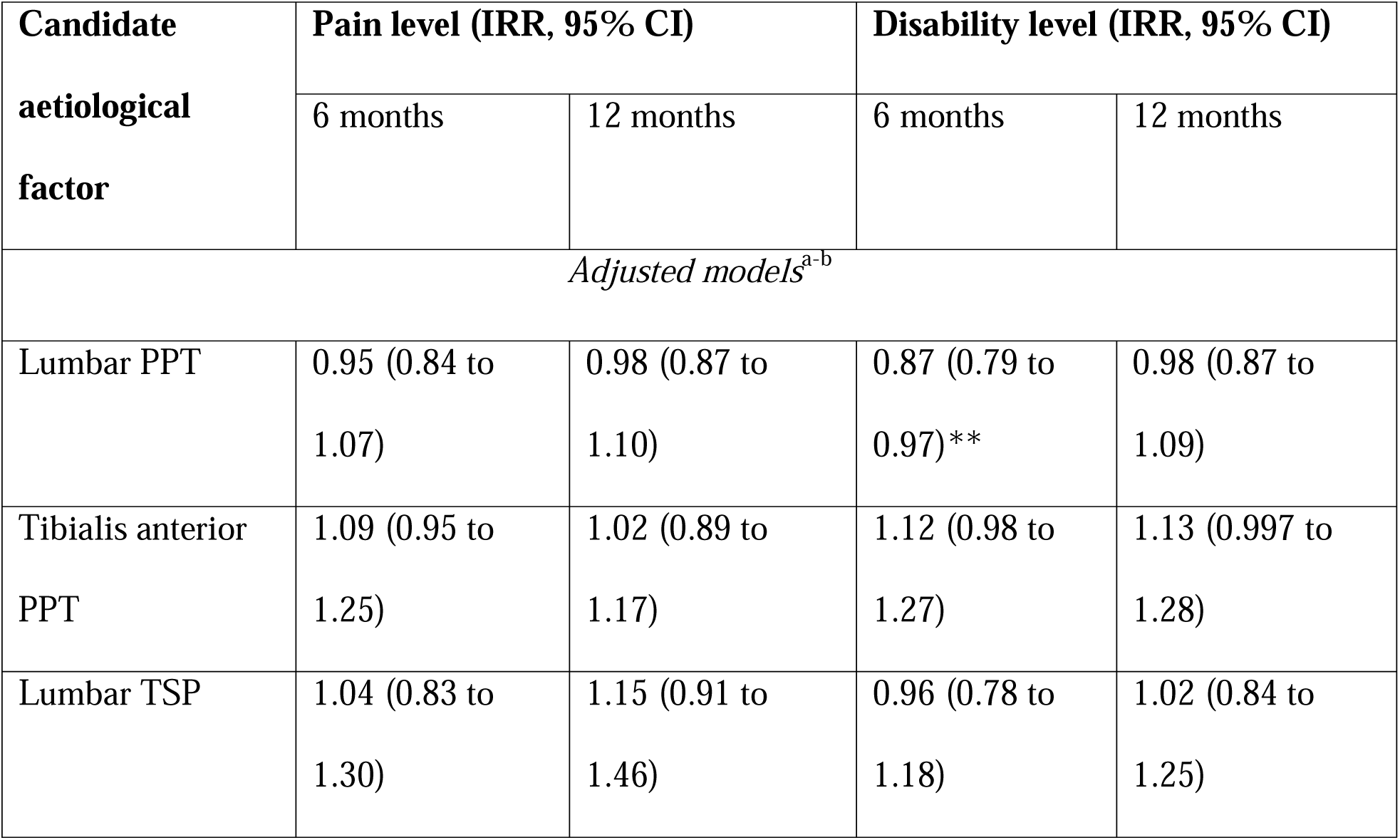
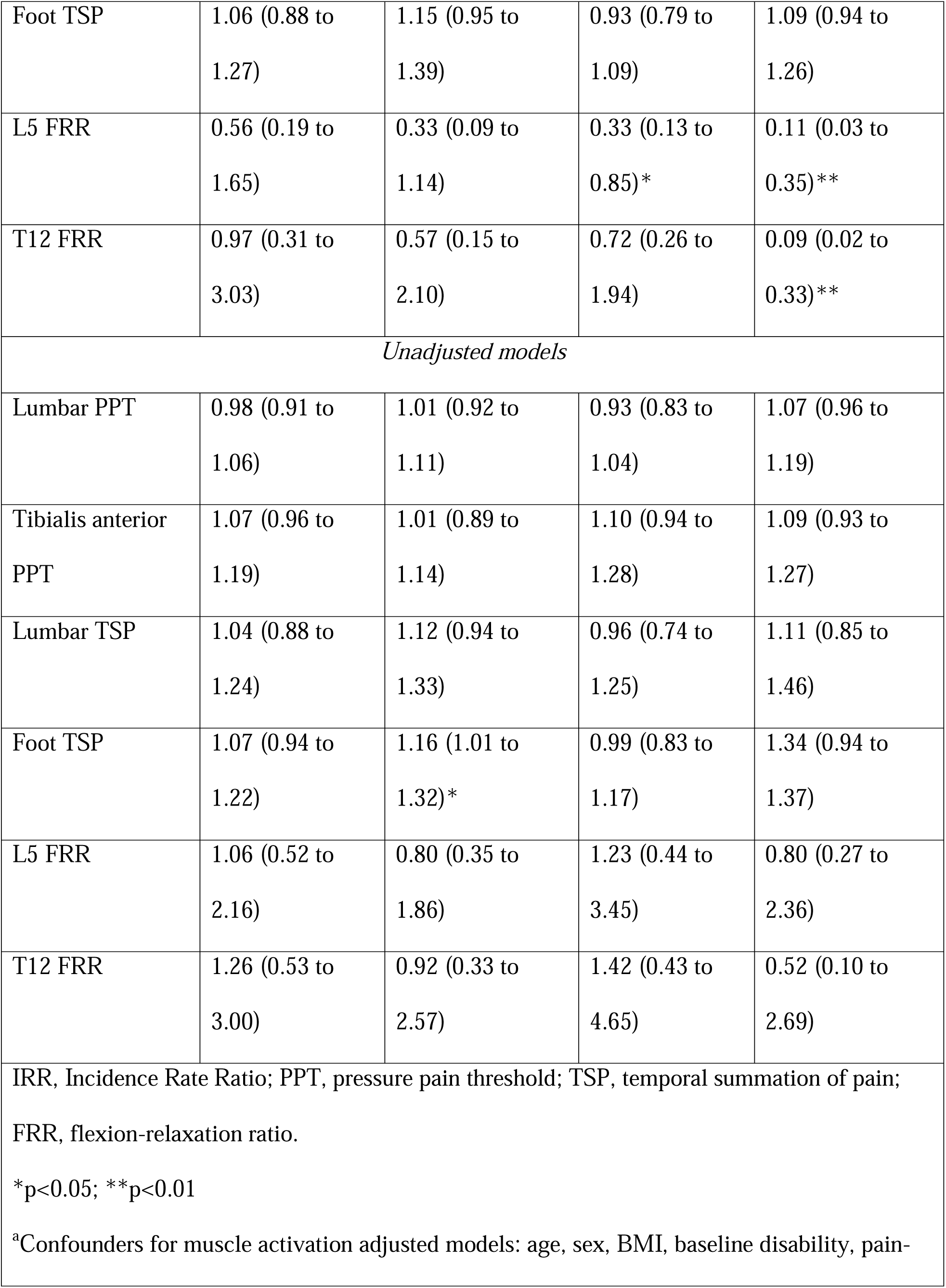
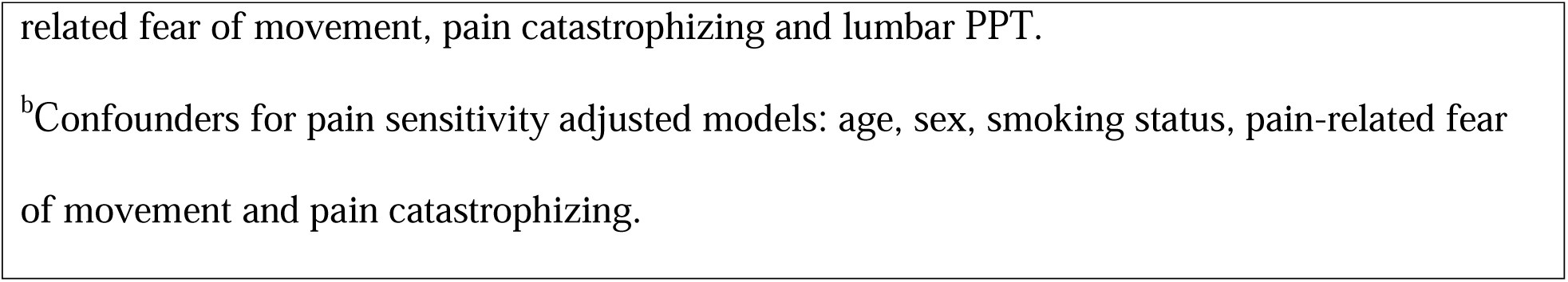
Negative binomial regression models to test the effect of pain sensitivity and muscle activity exposures on pain and disability levels at 6- and 12-months follow-ups.

| Candidate<br>aetiological<br>factor | Pain level (IRR, 95% CI) |  | Disability level (IRR, 95% CI) |  |
| --- | --- | --- | --- | --- |
|  | 6 months | 12 months | 6 months | 12 months |
| *Adjusted models<sup>a-b</sup>* |  |  |  |  |
| Lumbar PPT | 0.95 (0.84 to 1.07) | 0.98 (0.87 to 1.10) | 0.87 (0.79 to 0.97)** | 0.98 (0.87 to 1.09) |
| Tibialis anterior PPT | 1.09 (0.95 to 1.25) | 1.02 (0.89 to 1.17) | 1.12 (0.98 to 1.27) | 1.13 (0.997 to 1.28) |
| Lumbar TSP | 1.04 (0.83 to 1.30) | 1.15 (0.91 to 1.46) | 0.96 (0.78 to 1.18) | 1.02 (0.84 to 1.25) |
| Foot TSP | 1.06 (0.88 to 1.27) | 1.15 (0.95 to 1.39) | 0.93 (0.79 to 1.09) | 1.09 (0.94 to 1.26) |
| L5 FRR | 0.56 (0.19 to 1.65) | 0.33 (0.09 to 1.14) | 0.33 (0.13 to 0.85)* | 0.11 (0.03 to 0.35)** |
| T12 FRR | 0.97 (0.31 to 3.03) | 0.57 (0.15 to 2.10) | 0.72 (0.26 to 1.94) | 0.09 (0.02 to 0.33)** |
| *Unadjusted models* |  |  |  |  |
| Lumbar PPT | 0.98 (0.91 to 1.06) | 1.01 (0.92 to 1.11) | 0.93 (0.83 to 1.04) | 1.07 (0.96 to 1.19) |
| Tibialis anterior PPT | 1.07 (0.96 to 1.19) | 1.01 (0.89 to 1.14) | 1.10 (0.94 to 1.28) | 1.09 (0.93 to 1.27) |
| Lumbar TSP | 1.04 (0.88 to 1.24) | 1.12 (0.94 to 1.33) | 0.96 (0.74 to 1.25) | 1.11 (0.85 to 1.46) |
| Foot TSP | 1.07 (0.94 to 1.22) | 1.16 (1.01 to 1.32)* | 0.99 (0.83 to 1.17) | 1.34 (0.94 to 1.37) |
| L5 FRR | 1.06 (0.52 to 2.16) | 0.80 (0.35 to 1.86) | 1.23 (0.44 to 3.45) | 0.80 (0.27 to 2.36) |
| T12 FRR | 1.26 (0.53 to 3.00) | 0.92 (0.33 to 2.57) | 1.42 (0.43 to 4.65) | 0.52 (0.10 to 2.69) |
| IRR, Incidence Rate Ratio; PPT, pressure pain threshold; TSP, temporal summation of pain; FRR, flexion-relaxation ratio.<br>*p<0.05; **p<0.01<br><sup>a</sup>Confounders for muscle activation adjusted models: age, sex, BMI, baseline disability, pain- |  |  |  |  |

related fear of movement, pain catastrophizing and lumbar PPT.
<sup>b</sup>Confounders for pain sensitivity adjusted models: age, sex, smoking status, pain-related fear of movement and pain catastrophizing.

## DISCUSSION

### Key results

In this causation analysis of longitudinal data, we found that higher erector spinae muscle activation in the acute phase of LBP was associated with lower levels of disability at both 6 and 12 months. In addition, lower lumbar PPTs were associated with higher levels of pain-related disability at 6 months. While the findings related to pain sensitivity are consistent with our initial hypothesis, those concerning muscle activity do not support it.

### Comparison with existing literature

#### Erector spinae muscle activation

The results do not support our initial hypothesis, but are in line with the “motor adaptation to pain” theory proposed by Hodges & Tucker in 2011, suggesting that early motor adaptations to pain could protect the painful region and facilitate healing.^24^ In our study, greater erector spinae muscle activation during a full trunk flexion explained lower levels of disability at both 6 and 12 months, which supports the benefits of short-term adaptations in motor control. Based on the latter theory, increased muscle activation in the acute phase may act as a protective role for the painful lumbar spine during end-range movement, thereby promoting recovery. Our initial hypothesis was informed by evidence of the persistence of greater muscle activation in individuals with CLBP. However, it remains unclear how erector spinae muscle activation fluctuates during the transition from acute to chronic pain. For instance, does muscle activation normalize during the acute or subacute phases (i.e., decrease from initial high levels), thus preventing potential long-term adverse effects such as increased spinal load and reduced movement or velocity)?^24^ Conversely, in individuals with lower muscle activation in the acute phase of LBP, does muscle activation increase in time? Addressing these questions will require longitudinal studies tracking fluctuation of erector spinae muscle activation across acute and subacute phases of LBP. Moreover, we might question why muscle activation in the acute phase predicted disability levels but not pain intensity in the long-term. A hypothesis could be that although NPRS, the scale used to measure pain intensity, has good to excellent test-retest reliability,^35^ its between-subject variability might have prevented to capture a clear trend in associations between sensorimotor outcomes in the acute stage of LBP and long-term pain intensity. Indeed, it was shown that for different individuals, the same scores on NPRS represented distinct sensations.^57^ On the opposite, ODI has multiple anchors for each domain assessed, which could reduce the variability between subjects with similar conditions. This might allow ODI to better discriminate between subjects with higher and lower disability levels and facilitate the examination of causal associations from exposures to outcomes.

An interesting parallel may be drawn between our results on motor exposures and the role of inflammatory processes in the recovery from ALBP. A longitudinal study conducted by Klyne et al. has shown that patients with ALBP who had recovered at 6 months initially had higher blood concentrations of C-reactive protein, a pro-inflammatory molecule, compared to those who did not recover.^58^ Likewise, Parisien et al. (2022) conducted a cohort study of patients with ALBP and observed that an early inflammatory response was associated with pain resolution.^59^

Together, these findings suggest that initial increase in erector spinae activation, like early inflammatory response, may represent adaptative and potentially beneficial mechanisms to foster healing in the acute phase of LBP. Clinically, this may suggest that an increase in erector spinae muscle activation during a trunk flexion in ALBP could be a beneficial protective reaction in the short-term. As such, attempts to normalize this response through targeted interventions may be unwarranted. Nonetheless, future studies with corrections for multiple analyses replicating these results are needed to confirm this recommendation.

#### Pain sensitivity

The results obtained regarding the association of a pain sensitivity measure (lumbar PPT) and the mean disability level at 6 months align with those from a meta-analysis by Georgopoulos et al., which reported that baseline mechanical thresholds such as PPT predict future disability in individuals with musculoskeletal pain.^23^ However, our results partially differ from previous cohort studies in which pain sensitivity in ALBP did not predict the transition to chronic pain^19^ or recovery.^20,21^ In the present study, only one of four pain sensitivity measures (lumbar PPT) was associated with long-term disability, and none was associated with pain. The fact that PPT over the low back area provides the biggest effect sizes between LBP participants and pain-free controls^17^ could partly explain why only this measure was significantly associated with disability.

Our results however cannot be directly compared with those of prior studies on the prediction of the transition to chronic pain,^19–21^ as the study designs differ greatly. First, most studies used dichotomic outcomes: recovered or not (chronic), whereas we examined continuous measures of pain and disability. Dichotomisation can introduce variability due to differing definitions of recovery or CLBP, whereas continuous outcomes avoid such arbitrary thresholds. Second, to our knowledge, this is the first study that controlled for a MSAS of potential confounders, identified using DAGs for pain sensitivity in ALBP. The need to control for confounding factors (e.g., variables that may influence both the exposure and outcome) in causation analyses distinguishes them from predictive studies, where this is not required, the aim being to accurately predict the outcome.^6^ Consequently, causal models are typically theory-driven, whereas prediction models are usually data-driven.^6^ These differences in study aims and analytical approach may explain why this study captured an association between a measure of HACS in ALBP and long-term disability levels, whereas previous studies did not. Our results suggest that HACS, measured locally to the painful region in ALBP, which could primarily, although not exclusively, reflect peripheral sensitization,^23,38^ could be a causal mechanism of the persistence of LBP. These results should however be interpreted with great caution as only one pain sensitivity model out of 16 reached statistical significance and this could be due to chance as we did not correct for multiple testing. Future well-powered studies should be conducted to confirm those results.

### Strengths and limitations

A major strength of this study is the use of DAGs to determine a sufficient set of confounders to control for in our statistical models. This approach provides a transparent and standardised method for confounders selection, which is crucial to accurately estimate a causal association between exposures and outcomes.^45^ To our knowledge, this is the first study to examine associations between erector spinae muscle activation and pain sensitivity in ALBP in relation to long-term pain and disability outcomes, while rigorously controlling for confounders. This study also presents limitations. First, as an exploratory analysis, no primary outcome was pre-specified, and multiple statistical analyses were conducted without correcting for multiplicity, which increases the risk of Type I error (false positive).^60^ Accordingly, the findings should be interpreted with caution. Second, the generalisability of the results may be limited to community-dwelling people with ALBP and may not extend to patients seeking care in clinical settings (e.g., emergency departments). Third, as the sensorimotor outcomes were solely measured at short-term, their trajectory in time remains unknown, limiting the understanding of their impact on the development of CLBP. For example, in those who had an increased muscle activation at baseline, a decrease in muscle activation in time would support that this increase was potentially protective.

### Conclusions

This exploratory study provides evidence that, in individuals with ALBP, higher erector spinae muscle activation is associated with lower levels of pain-related disability at 6 months and 12 months after an acute episode. This association suggests that increased erector spinae muscle activation in the acute phase of LBP may represent a protective and potentially beneficial mechanism to foster disability improvement. Moreover, higher lumbar pressure pain sensitivity at baseline was associated with disability at 6 months; however, this association was not consistently supported across other pain sensitivity measures. Longitudinal measurement of pain sensitivity and muscle activation are needed to better understand their trajectory in the acute and subacute phases of LBP and improve our understanding of their potential contribution in the development of chronic pain.

## Data Availability

All data produced in the present study are available upon reasonable request to the authors

## Declaration of AI usage

No Generative AI was used to create the content of this manuscript.

## Acknowledgements

We thank Eric McArthur, biostatistician, for his help in statistical analysis, and Alexandre Desgagné-Lebeuf, engineer, for the treatment of EMG data.

## Data Statement

Data are available upon reasonable request to authors.

