## Supplementary file 1 for "Could sensorimotor factors in acute low back pain explain long-term pain and disability? A secondary analysis of longitudinal data from a randomised controlled trial"

**Supplementary file 1.** Codes used to build the Directed Acyclic Graphs (DAGs)

*A. Muscle activation DAG*

dag {

bb="-4.086,-4.623,4.671,3.658"

"Baseline disability" [pos="3.004,1.512"]

"Long-term pain" [outcome,pos="3.500,-2.239"]

"Muscle activation" [exposure,pos="1.424,2.243"]

"Pain sensitivity" [pos="2.175,-3.860"]

"Pain-related fear" [pos="-0.011,2.339"]

"Self-efficacy" [pos="-1.480,1.926"]

Age [pos="-2.297,-0.125"]

BMI [pos="3.644,-0.490"]

Catastrophizing [pos="-2.286,-1.969"]

Sex [pos="-1.524,-3.129"]

Smoking [pos="-0.144,-4.083"]

"Baseline disability" -> "Long-term pain"

"Baseline disability" -> "Muscle activation"

"Muscle activation" -> "Long-term pain"

"Pain sensitivity" -> "Baseline disability"

"Pain sensitivity" -> "Long-term pain"

"Pain sensitivity" -> "Muscle activation"

"Pain-related fear" -> "Long-term pain"

"Pain-related fear" -> "Muscle activation"

"Pain-related fear" -> "Pain sensitivity"

"Self-efficacy" -> "Baseline disability"

"Self-efficacy" -> "Long-term pain"

Age -> "Baseline disability"

Age -> "Long-term pain"

Age -> "Muscle activation"

Age -> "Pain sensitivity"

Age -> "Pain-related fear"

Age -> "Self-efficacy"

Age -> BMI

Age -> Catastrophizing

Age -> Smoking

BMI -> "Baseline disability"

BMI -> "Long-term pain"

BMI -> "Muscle activation"

Catastrophizing -> "Long-term pain"

Catastrophizing -> "Muscle activation"

Catastrophizing -> "Pain sensitivity"

Catastrophizing -> "Pain-related fear"

Sex -> "Long-term pain"

Sex -> "Muscle activation"

Sex -> "Pain sensitivity"

Sex -> BMI

Sex -> Smoking

Smoking -> "Baseline disability"

Smoking -> "Long-term pain"

Smoking -> "Pain sensitivity"

Smoking -> BMI

}

*B. Pain sensitivity DAG*

dag {

bb="-4.086,-4.623,4.671,3.658"

"Baseline disability" [pos="3.004,1.512"]

"Long-term pain" [outcome,pos="3.500,-2.239"]

"Pain sensitivity" [exposure,pos="2.175,-3.860"]

"Pain-related fear" [pos="1.469,2.529"]

"Self-efficacy" [pos="-0.939,1.957"]

Age [pos="-2.297,-0.125"]

BMI [pos="3.644,-0.490"]

Catastrophizing [pos="-2.286,-1.969"]

Sex [pos="-1.524,-3.129"]

Smoking [pos="-0.144,-4.083"]

"Baseline disability" -> "Long-term pain"

"Pain sensitivity" -> "Baseline disability"

"Pain sensitivity" -> "Long-term pain"

"Pain-related fear" -> "Long-term pain"

"Pain-related fear" -> "Pain sensitivity"

"Self-efficacy" -> "Baseline disability"

"Self-efficacy" -> "Long-term pain"

Age -> "Baseline disability"

Age -> "Long-term pain"

Age -> "Pain sensitivity"

Age -> "Pain-related fear"

Age -> "Self-efficacy"

Age -> BMI

Age -> Catastrophizing

Age -> Smoking

BMI -> "Baseline disability"

BMI -> "Long-term pain"

Catastrophizing -> "Long-term pain"

Catastrophizing -> "Pain sensitivity"

Catastrophizing -> "Pain-related fear"

Sex -> "Long-term pain"

Sex -> "Pain sensitivity"

Sex -> BMI

Sex -> Smoking

Smoking -> "Baseline disability"

Smoking -> "Long-term pain"

Smoking -> "Pain sensitivity"

Smoking -> BMI

}
