## Supplementary file 2 for "Could sensorimotor factors in acute low back pain explain long-term pain and disability? A secondary analysis of longitudinal data from a randomised controlled trial"

**Supplementary file 2.** Relationships between variables and references used to build directed acyclic graphs

| **Edge** | **Study design** | **References** |
| --- | --- | --- |
| Age 🡪 Pain | *Expert opinion* based on a topical review | Mullins 2022 (1) |
| Age 🡪 Muscle activation | Cross-sectional associations | Zhang 2024 (2) |
| Age 🡪 Catastrophizing | Cross-sectional associations | Murray 2021 (3) |
| Age 🡪 Pain-related fear | *Expert opinion* based on a critical literature review | Vlaeyen 2000 (4) |
| Age 🡪 Pain sensitivity | Systematic review and meta-analysis | El Tumi 2017 (5) |
| Age 🡪 Smoking | Cross-sectional associations | Kranjac 2025 (6) |
| Age 🡪 Baseline disability | Cross-sectional associations | Murray 2021 (3) |
| Age 🡪 Self-efficacy | Cross-sectional associations | Murray 2021 (3) |
| Age 🡪 BMI | Cross-sectional analysis of four longitudinal cohort studies | Yang 2021 (7) |
| Baseline disability 🡪 Pain | **Predictive model**  Longitudinal observational study. | Stevans 2021 (8) |
| Baseline disability 🡪 Muscle activation | **Cross-sectional associations**  **State-of-the-art clinical commentary** | Alschuler 2009 (9)  Hodges 2019 (10) |
| BMI 🡪 Baseline disability | **Cross-sectional analysis** of a Chronic pain patients’ cohort | Basem 2021 (11) |
| BMI 🡪 Pain | **Causal model** Cross-sectional analysis of a longitudinal observational study (Design-based confounding control)  **Cross-sectional analysis** of a Chronic pain patients’ cohort | Zheng 2025 (12)  Basem 2021 (11) |
| BMI – Pain-related fear | **Saturated** | |
| BMI – Self-efficacy | **Saturated** | |
| BMI – Catastrophizing | **Saturated** | |
| BMI 🡪 Muscle activation | *Expert opinion* based on a **cross-sectional study** of obesity and spinal ROM | Vismara 2010 (13) |
| BMI ⮾ Pain sensitivity | Cross-sectional analysis of healthy weight and obese participants found no difference in pain sensitivity between healthy weight and obese participants | Emerson 2021 (14) |
| Catastrophizing 🡪 Muscle activation | Cross-sectional associations | Henchoz 2013 (15) |
| Catastrophizing ⮾ Baseline disability | Systematic review and meta-analysis | Lee 2015 (16) |
| Catastrophizing 🡪 Pain-related fear | Critical literature review | Vlaeyen 2000 (4) |
| Catastrophizing 🡪 Pain | Systematic review | Wertli 2014 (17) |
| Catastrophizing 🡪 pain sensitivity | Cross-sectional mediation analysis | Meints 2020 |
| Catastrophizing – Self-efficacy | **Saturated** | |
| Pain-related fear 🡪 Pain | Systematic review and meta-analysis | Lee 2015 (16) |
| Pain-related fear 🡪 Pain sensitivity | Cross-sectional associations | Ismail 2025 |
| Pain-related fear 🡪 Muscle activation | Cross-sectional associations | Alschuler 2009 (9) |
| Pain-related fear – Self-efficacy | **Saturated** | |
| Pain sensitivity 🡪 Muscle activation | State-of-the-art clinical commentary | Hodges 2019 (10) |
| Pain sensitivity 🡪 Pain | Prospective longitudinal cohort study | Marcuzzi 2018 (18) |
| Pain sensitivity 🡪 Baseline disability | Systematic review and meta-analysis | Georgopoulos 2019 (19) |
| Self-efficacy 🡪 Baseline disability | Systematic review and meta-analysis | Lee 2015 (16) |
| Self-efficacy 🡪 Pain | Meta-analysis | Jackson 2014 (20) |
| Self-efficacy ⮾ Pain sensitivity | No evidence | |
| Sex – Baseline disability | **Saturated** | |
| Sex 🡪 BMI | There is a higher prevalence of severe and morbid obesity in women than in men.  **Pooled 1698 population-based data sources** | Di Cesare 2016 (NCD Risk Factor Collaboration) |
| Sex – Catastrophizing | **Saturated** | |
| Sex 🡪 Pain | Literature review | Smith 2025 (21) |
| Sex 🡪 Muscle activation | Cross-sectional associations | Kienbacher 2015 (22) |
| Sex 🡪 Pain sensitivity | Cross-sectional associations | Vogel 2023 (23) |
| Sex ⮾ Self-efficacy | **Cross-sectional associations study** showed no effect of sex on self-efficacy | Chong 2001 (24) |
| Sex 🡪 Smoking | **Cross-sectional associations** | Kranjac 2025 (6) |
| Smoking🡪 Pain | **Predictive model**  Longitudinal observational study | Stevans 2021 (8) |
| Smoking – Catastrophizing | **Saturated** | |
| Smoking – Self-efficacy | **Saturated** | |
| Smoking – Pain-related fear | **Saturated** | |
| Smoking 🡪 BMI | **Meta-analysis** | Whitlock 2009 (25) |
| Smoking 🡪 Pain sensitivity | **Cross-sectional associations study** | Chiba 2024 (26) |
| Pain is the long-term outcome (6 and 12 months), including pain intensity and pain-related disability  In blue: We did not include this arrow as the MSAS was saturated, i.e. the addition of a given arrow did not change the MSAS. | | |
