## Supplementary file 3 for "Could sensorimotor factors in acute low back pain explain long-term pain and disability? A secondary analysis of longitudinal data from a randomised controlled trial"

**Supplementary file 3.** Pearson correlation coefficients of covariates with outcomes

| **Variables** | **Pain level** | | **Disability level** | |
| --- | --- | --- | --- | --- |
|  | *6 months* | *12 months* | *6 months* | *12 months* |
| Sex | 0.14 | 0.09 | 0.15 | 0.09 |
| Age | 0.05 | 0.23* | 0.11 | 0.22* |
| Smoking status | -0.13 | -0.05 | -0.10 | -0.2 |
| BMI | 0.27* | 0.07 | 0.45** | 0.16 |
| Disability^a^ | 0.22* | 0.13 | 0.27* | 0.22* |
| Kinesiophobia | 0.14 | 0.10 | 0.16 | 0.23* |
| Pain catastrophizing | 0.28** | 0.23* | 0.26* | 0.22* |
| Self-efficacy | -0.20 | -0.15 | -0.20 | -0.08 |
| BMI, body mass index.  ^a^Baseline disability  *p<0.05; **p<0.01 | | | | |
